# Sensitive Glioma Detection and Recurrence Monitoring Using a Machine Learning Model Based on Circulating Monocytes

**DOI:** 10.64898/2026.05.29.26354409

**Authors:** Wei Wu, Ruichao Chai, Peng Xia, Lingxiang Wu, Benqing Yu, Xiaoyong Chen, Bo Pang, Di Chen, Yu Wang, Na Wang, Xuejun Li, Hongwei Liu, Qiwen Deng, Feng Wan, Feng Lyu, Liang Wang, Wei Zhang, Junxia Zhang, Tao Jiang, Qianghu Wang

## Abstract

**Background:** Glioma induces profound systemic immune alterations despite its anatomical confinement to the central nervous system. Circulating immune cells, particularly monocytes, are key mediators of tumor-host crosstalk and may retain tumor-induced transcriptional imprints. However, their potential clinical utility as blood-based biomarkers for detection and monitoring, remain largely unexplored.

**Methods and findings:** In this study, we performed integrated single-cell RNA sequencing of blood immune cells and demonstrated that circulating CD14^+^ monocytes are significantly expanded in glioma patients, exhibiting features of differentiation arrest and increased transcriptional plasticity. These cells harbor glioma-specific molecular signatures distinct from those observed in healthy controls and patients with other tumors. Leveraging these findings, we developed an ensemble machine learning diagnostic model based on transcriptomic profiles of circulating CD14^+^ monocytes (training cohort, n = 107), which achieved a mean area under the receiver operating characteristic curve (AUC) of 0.975 during cross-validation. In an independent cohort of 567 participants, the model maintained high diagnostic accuracy, yielding an AUC of 0.888 for distinguishing glioma from controls and other tumors. And it achieved a recurrence detection AUC of 0.975 in 51 postoperative samples. Moreover, in a follow-up study involving 30 glioma patients, lower model-derived scores of postoperation were significantly associated with prolonged progression-free survival (log-rank test, *P* = 0.034), supporting its prognostic utility.

**Conclusion:** We demonstrate circulating CD14^+^ monocytes undergo glioma-specific transcriptional reprogramming, generating systemic tumor-associated signal captured via transcriptomic profiling. This blood-based diagnostic model provides non-invasive, scalable approach for glioma detection, recurrence surveillance, outcome prediction.

**Author summary:** *Why was this study done?:*

- Diagnosis and recurrence monitoring for glioma remain challenging with current MRI and biopsy.
- Gliomas alter systemic immunity, but whether circulating monocytes carry tumor-specific signals remains unclear.
- We aimed to develop a blood-based test using circulating monocyte transcriptomes for glioma detection and monitoring.

*What did the researchers do and find?:*

- Single-cell RNA-seq revealed that glioma patients have more CD14 monocytes with abnormal differentiation.
- We built a machine learning model based on monocyte gene expression. It achieved a diagnostic AUC of 0.888 in 567 independent samples and a recurrence detection AUC of 0.975 in 51 postoperative samples.
- In a follow-up study of 30 patients, lower postoperative model-derived scores predicted longer progression-free survival (*P* = 0.034).

*What do these findings mean?:*

- Circulating monocytes capture glioma-specific transcriptional reprogramming features, enabling non-invasive liquid biopsy.
- The model may help distinguish recurrence from pseudo-progression and guide postoperative risk stratification.
- Larger prospective multicenter validation studies are needed to further confirm clinical generalizability.

## Introduction

Adult-type diffused gliomas, including isocitrate dehydrogenase (IDH)-mutant astrocytoma, IDH-mutant and 1p/19q co-deleted oligodendrocytes, and IDH-wildtype GBM, are the most common primary malignant brain tumor in the Central Nervous System (CNS) [1–3]. While magnetic resonance imaging (MRI) and histopathological analysis of tissue biopsies remain the cornerstone of tumor diagnosis, these conventional approaches cannot fulfill the clinical needs of gliomas. A portion of tumors demonstrate subtle imaging features that challenge definitive characterization, particularly in cases of some IDH-mutant gliomas [4, 5]. In addition, conventional MRI faces a diagnostic conundrum in distinguishing true tumor progression from treatment-induced pseudo progression, particularly in IDH-mutant gliomas, where the incidence of pseudo progression can reach up to 39% [6, 7]. This diagnostic ambiguity remains even when employing advanced perfusion imaging metrics [8]. Concurrently, stereotactic biopsy procedures are associated with a hemorrhage risk and often suffer from considerable spatial heterogeneity [9, 10]. Genomic analyses further reveal up to 68% discordance between biopsy and resection specimens in multifocal glioblastomas [11–13].

While gliomas actively secrete molecular cues into the systemic circulation, including tumor-derived biomarkers such as proteins, and circulating tumor cells (CTCs) [14], their detectability in peripheral blood during early disease stages remains limited by the blood-brain barrier (BBB). The selective interface property of BBB exerts physical and functional constraints on the transfer of macromolecular biomarkers from the tumor microenvironment to the systemic circulation, thereby reducing their bioavailability in peripheral blood during glioma development [15, 16].

The interplay between tumor cells and diverse immune cell populations derived from peripheral circulation may present a novel avenue for inferring glioma status. In particular, the circulating monocytes are recruited from the peripheral blood into the glioma microenvironment, where glioma-derived signals induce their polarization toward an immunosuppressive phenotype, thereby facilitating tumor evolution [17]. At the cellular level, single-cell transcriptomic analyses have elucidated the functional heterogeneity of monocyte-derived tumor associated macrophages (TAMs). The peri-necrotic niche-resident monocyte-derived TAMs exhibiting remarkable hypoxic and inflammatory transcriptional signature [18] and tumor-associated monocytes with immunosuppression and high expression of AREG and EREG genes [19] have been specifically revealed in glioma patients. These findings illuminate the dynamic interplay between systemic immunity and glioma progression, highlighting the capacity of gliomas to exploit peripheral myeloid populations for local immune modulation and malignant expansion. Within this context, circulating monocytes may undergo functional reprogramming or conditioning prior to crossing the BBB, leaving detectable transcriptional signatures in the blood.

In this study, we performed integrated bulk and single-cell RNA sequencing (scRNA-seq) of circulating CD14^+^ monocytes from glioma patients, comparing them to healthy controls (HCs) and patients with other solid tumors. Based on the transcriptional profiles of circulating CD14^+^ monocytes from 107 glioma patients (training cohort), we developed a diagnostic model using an ensemble machine learning approach. The model achieved a mean cross validation AUC of 0.975 for distinguish glioma patients from other samples. In an independent validation cohort of 567 participants, it maintained robust performance, yielding an AUC of 0.888 for distinguishing glioma from controls and other tumors. And it achieved an AUC of 0.975 for detecting recurrence within two years in 51 postoperative samples. Furthermore, in a follow up study of 30 glioma patients, lower postoperative model-derived scores were significantly associated with prolonged progression free survival (PFS) (log rank test, *P* = 0.034). Collectively, our findings demonstrate that transcriptional profiling of circulating CD14^+^ monocytes enables non invasive glioma detection, recurrence surveillance, and prognostic assessment.

## Results

### Proportion of Circulating CD14^+^ monocytes were elevated in glioma patients

To investigate the impact of tumor on systemic immune responses, we integrated single-cell transcriptional profiling of peripheral immune cells from glioma (n = 9), head and neck squamous cell carcinoma (HNSCC, n = 26), non-small cell lung cancer (NSCLC, n = 33), and healthy controls (HC, n = 16) (**Figure 1A**). We isolated 343,773 immune cells defined by CD45 expression levels exceeding a threshold (cutoff = 1) and clustered these cells into 16 distinct cell clusters (**Figures 1A; Supplementary Figure S1A**). Subsequent lineage-specific marker annotation defined these cell clusters into eight major cell types (**Figures 1B, C; Supplementary Figure S1B**), including T/NK cells (n = 119,331) marked by NKG7, KLRD1 and CST7, T cells (n = 105,717) which expressed CD2, CD3D and CD3E, B cells (n = 18,439) marked by CD40, MS4A1 and CD79A, plasma cells (n = 558) identified by the expression of MZB1, CD14 monocytes (n = 81,332) marked by CD14 and S100A9, CD16 monocytes (n = 12,212) marked by FCGR3A and MS4A7, dendritic cells (n = 4,164) identified by the expression of FCGR1A and CST3, and platelets (n = 2,020) which expressed PPBP and PF4. In addition, there were 6,948 cells that were not defined due to the lack of clear lineage-specific markers (**Supplementary Figure S1B**).

**Figure 1.**
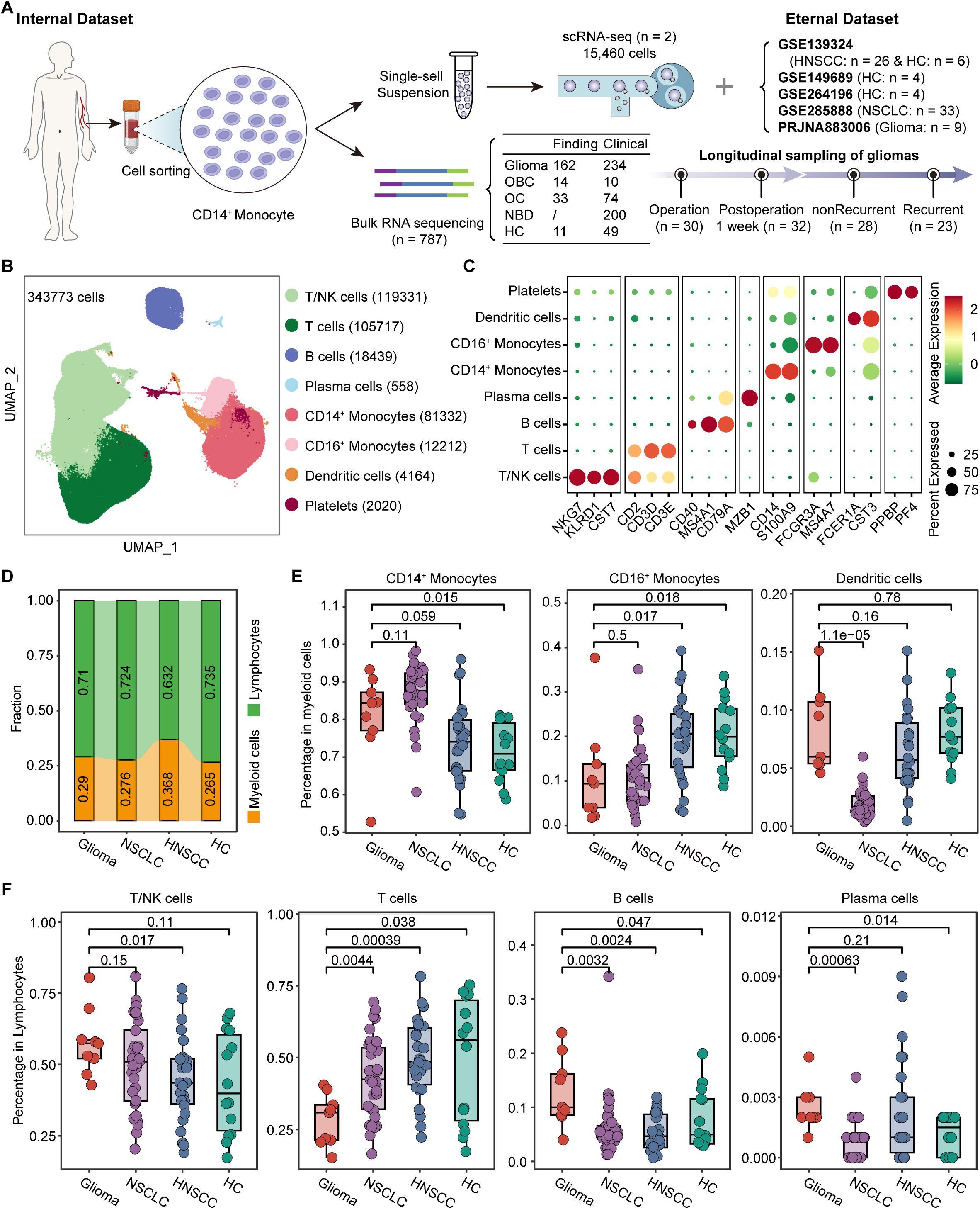
Comparison of the composition of peripheral immune cells among different types of samples. **(A)** Schematic overview of the experimental workflow. OBC, other brain tumors; OC, non-central nervous system cancers; HNSCC, head and neck squamous cell carcinoma; NSCLC, non-small cell lung cancer; NBD, non-neoplastic brain disorder; HC, healthy controls. **(B)** UMAP plot showing major 8 cell types of 82 samples identified by integrated analysis. Each dot corresponds to a single cell, colored by cell types. NK cells, natural killer cells. **(C)** Dot plot showing average expression of known markers in indicated cell types. The dot size represents percent of cells expressing the genes in each cluster. The expression intensity of markers is shown by color. **(D)** The stacked bar plot showing the proportions of peripheral immune cells, including Lymphocytes and myeloid cells, in different types of samples. (**E, F**) Comparison of the cell abundances of 3 types of myeloid cells (**E**) and 4 types of Lymphocytes (**F**). P value was obtained by the Wilcoxon rank-sum test.

We quantified the relative proportions of lymphoid and myeloid lineage cells across different sample types and found no major variation in lineage-level distribution between glioma patients and HCs, nor between glioma and other cancer types (**Figure 1D**). We subsequently investigated the intra-lineage composition of myeloid cells as illustrated in **Figure 1E**, our analysis revealed a significant enrichment of CD14^+^ monocytes in both glioma and NSCLC patients compared to HCs (Wilcoxon rank-sum test, *P* < 0.05). However, no statistically significant differences were observed between these two cancer types regarding the abundance of CD14^+^ monocytes (**Figure 1E**). Within the lymphoid lineage, the proportion of T cells showed marked decrease in gliomas compared to other groups (Wilcoxon rank-sum test, *P* < 0.05) **(Figure 1F**). Conversely, B cells (Wilcoxon rank-sum test, *P* < 0.05) and plasma cells (Wilcoxon rank-sum test, *P* < 0.05) were more abundant in gliomas compared to HCs and other two cancer types, although their overall abundance remained low (**Figure 1F**). Collectively, these findings collectively highlight lineage-specific immune dysregulation patterns across different cancer types, with myeloid and lymphoid compartments exhibiting distinct alterations across glioma.

### Circulating CD14^+^ monocytes exhibit abnormal differentiation status in glioma patients

Given the high proportion of CD14^+^ monocytes observed in gliomas, we further characterized their functional properties by isolating CD14^+^ monocytes from two glioma blood samples through cell sorting (**Figure 1A**). These isolated cells were subjected to droplet-based scRNA-seq to profile their gene expression (detailed in the Methods), following strict quality control procedures, a total of 15,460 cells were retained for analysis (**Supplementary Figures S2A-C**). We then extracted 23,912 circulating CD14^+^ monocytes from public datasets (9 glioma patients and 14 HCs) and 13,469 circulating CD14^+^ monocytes from our own dataset (2 glioma patients), and reclustered these CD14^+^ monocytes to identify four transcriptionally distinct subclusters (**Figure 2A**). Proportional analysis revealed partial differences in the relative abundance of these four subclusters between glioma patients and HCs (**Figure 2B**). Differential expression gene (DEG) analysis was performed for each subcluster, revealing varying degrees of transcriptional dysregulation among glioma patients across all four subclusters (**Supplementary Table S3 and Figure 2C**). The upregulated genes in each subcluster were enriched in multiple tumor-associated KEGG pathways, including “Transcriptional mis-regulation in cancer”, “PI3K-Akt signaling pathway”, and “Toll-like receptor signaling pathway”, highlighting the potential involvement of circulating CD14^+^ monocytes in tumor-promoting immune modulation and glioma pathophysiology (**Figure 2C)**.

**Figure 2.**
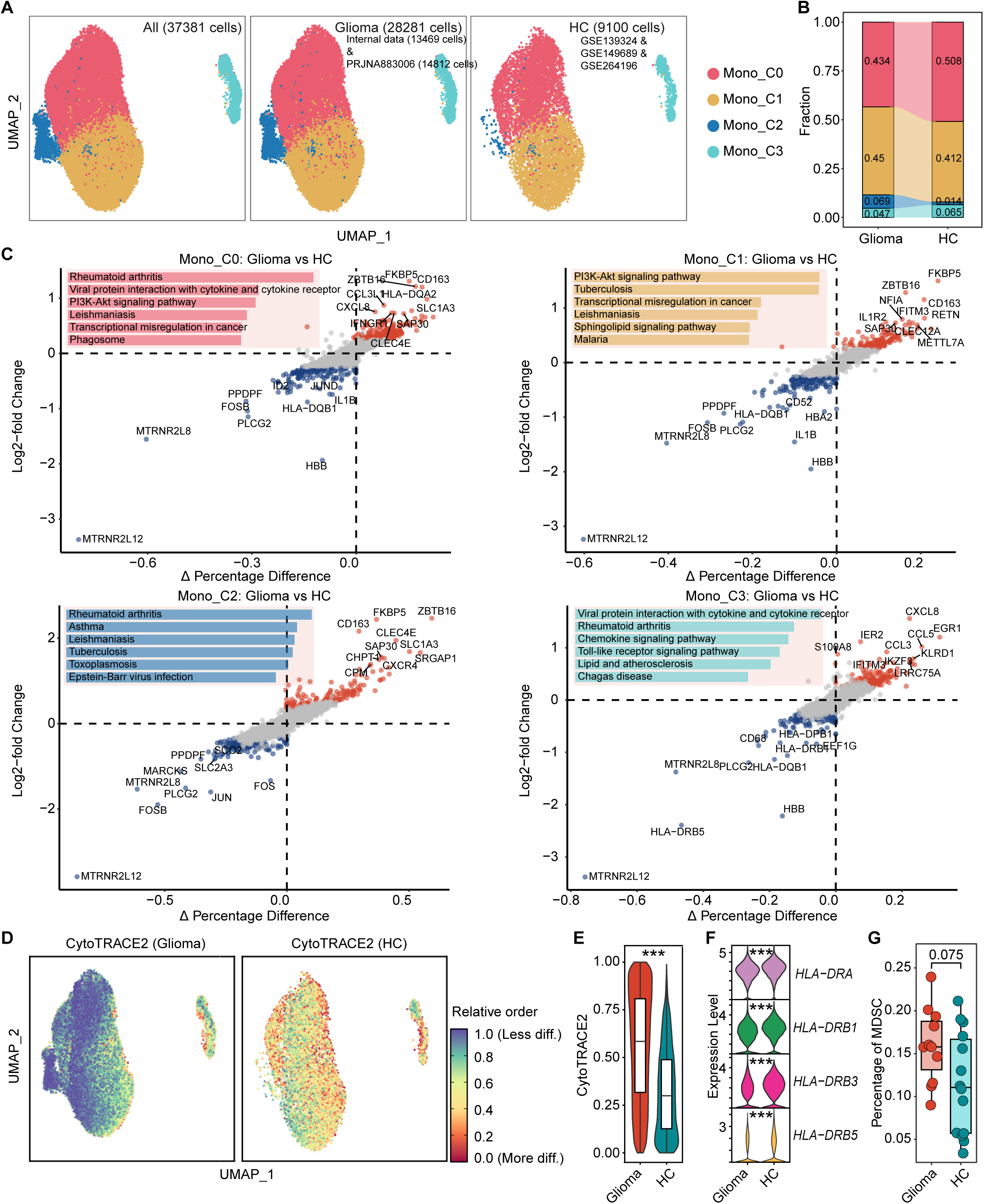
Characterization of peripheral CD14^+^ monocytes clusters between gliomas and HCs. **(A)** UMAP plot of 35511 peripheral CD14^+^ monocytes grouped into 4 cell subclusters from 9 glioma patients and 14 HCs (left, all cells from glioma and HC samples; middle, cells from glioma samples; right, cells from HC samples). **(B)** The stacked bar plot showing the proportions of 4 CD14^+^ monocytes subclusters in different types of samples. **(C)** Differential gene expression analysis of four CD14^+^ monocyte subclusters between glioma samples and HCs. The x-axis represents the difference in the percentage of cells expressing each gene, while the y-axis indicates the log2 fold change in expression. Colored DEGs meet the criteria of an adjusted p-value < 0.05 and an average log2 fold change > 0.25. Top five KEGG pathways enriched for upregulated genes in each subcluster are displayed in the top-left corner. (**D, E**) UMAP plot depicting the distribution of CytoTRACE2 scores among CD14^+^ monocytes between glioma samples and HCs. Lower CytoTRACE2 scores (closer to 0.0) indicate a higher degree of differentiation, while higher scores (closer to 1.0) reflect a lower degree of differentiation (**D**). Boxplot compares the CytoTRACE2 scores between two groups (**E**). P value was obtained by the Wilcoxon rank-sum test; ***, p < 0.001; **, p < 0.01; *, p < 0.05; ns, p _≥_ 0.05 (P values also apply to F). **(F)** Violin plot of selected four canonical markers of monocyte differentiation and maturation showing normalized expression in CD14^+^ monocytes. **(G)** Comparison of MDSC proportions among CD14^+^ monocytes between glioma samples and HCs. P value was obtained by the one-sided Wilcoxon rank-sum test.

Additionally, we employed CytoTRACE2 [20] to evaluate the differentiation potential of circulating CD14^+^ monocytes obtained from glioma patients compared to those from HCs. The results indicated a notable global shift toward lower differentiation states in circulating CD14^+^ monocytes derived from glioma patients, as evidenced by significantly higher CytoTRACE2 scores (**Figures 2D, E)**. The expression levels of canonical markers (e.g., HLA-DRA, etc.) associated with monocyte differentiation and maturation were significantly reduced in circulating CD14^+^ monocytes derived from glioma patients (**Figure 2F)**, further supporting their diminished differentiation status. Collectively, these findings suggested enhanced transcriptional plasticity and a less differentiated phenotype among glioma-derived circulating CD14^+^ monocytes.

Furthermore, based on previous studies [21], we defined myeloid-derived suppressor cells (MDSCs) as cells that express CD14, CD33, and ITGAM while lacking FUT4 expression. Using this definition, we observed an increased proportion of MDSC-like monocytes in the peripheral blood of glioma patients compared with healthy controls (Wilcoxon rank-sum test, *P* = 0.075; **Figure 2G**). Although this difference did not reach conventional statistical significance, the trend suggests a potential enrichment of immunosuppressive monocyte populations in glioma, which may contribute to global immune regulation [21].

### Systematic transcriptional dysregulation of circulating CD14^+^ monocytes in glioma

Given the systemic abnormal differentiation observed in circulating CD14^+^ monocytes of glioma patients, we subsequently conducted RNA sequencing on these monocytes (**Figure 3A**), isolated through magnetic-activated cell sorting (MACS), from 209 patient samples and eleven HCs (**Supplementary Table S1**) to further validate this alteration. These 209 patient-derived samples comprised 79 primary glioma samples; 32 samples collected one week after surgical resection, including 30 samples paired with the corresponding intraoperative samples and 2 unpaired samples; 23 recurrent glioma samples; 28 samples from patients who remained recurrence-free for 1-2 years after surgery; 14 samples from patients with other brain tumors (OBC); and 33 samples from patients with other non-central nervous system cancers (OC). non-central nervous system cancersWe first compared 79 treatment-naïve primary glioma patients with eleven HCs. The t-distributed stochastic neighbor embedding (t-SNE) analysis based on the 6,000 most variable genes (selected by median absolute deviation, MAD) revealed a clear separation between gliomas and HCs (**Figure 3B**). Gene set enrichment analysis (GSEA) revealed significant enrichment of six cancer hallmarks in circulating CD14^+^ monocytes from gliomas compared to HCs, including ‘Allograft Rejection’ (*FDR* < 0.001), ‘Epithelial Mesenchymal Transition’ (*FDR* = 0), ‘Il2 Stat5 Signaling’ (*FDR* = 0.02), ‘Complement’ (*FDR* = 0.02), ‘Kras Signaling Dn’ (*FDR* = 0.07) and ‘Inflammatory Response’ (*FDR* = 0.07) (**Figure 3C**), which involves in regulating immune activation, cell migration, and inflammatory signaling. Using the edgeR package, we identified 233 significantly upregulated and 25 significantly downregulated DEGs in glioma-derived circulating CD14^+^ monocytes (**Supplementary Table S4 and Supplementary Figures S3A, B**). Pathway enrichment analysis of the 233 upregulated DEGs revealed significant associations with KEGG pathways functionally relevant to monocyte biology, including ‘Cytokine-cytokine receptor interaction’, ‘Transcriptional misregulation in cancer’, and ‘TGF-beta signaling pathway’ (**Figure 3D**), suggesting a strengthened immune communication in the circulating CD14^+^ monocytes.

**Figure 3.**
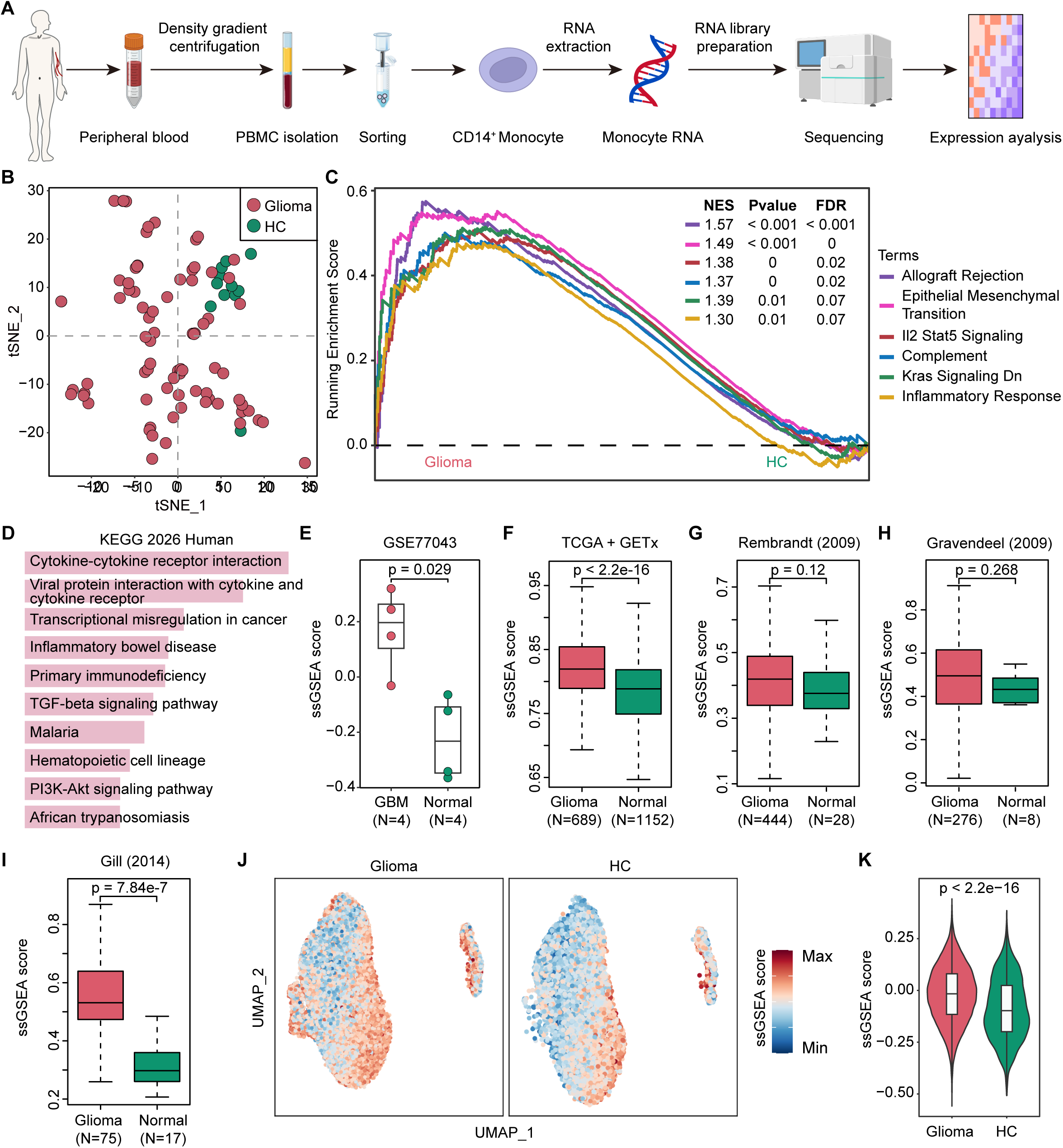
Bulk RNA-seq profiling of circulating CD14^+^ monocytes from glioma samples and HCs. **(A)** Schematic overview of the RNA sequencing workflow of CD14^+^ monocyte. **(B)** t-SNE plot showing the result of unsupervised dimensionality reduction on 79 glioma samples and 11 HCs using the top 6,000 highly variable genes. **(C)** Gene Set Enrichment Analysis (GSEA) of cancer hallmark pathways was performed to compare glioma samples and HCs. Pathways with positive NES are enriched in glioma samples. FDR: false discovery rate; NES: normalized enrichment score. **(D)** KEGG enrichment analysis of 233 upregulated DEGs in glioma samples. **(E)** Single sample GSEA (ssGSEA) comparison of the 233 DEGs using monocyte expression profiles from four glioma and four normal samples in the GSE77043 dataset. P value was obtained by the Wilcoxon rank-sum test (P values also apply to **G** and **I**). (**F-I**) ssGSEA comparison of the 233 DEGs using expression profiles from glioma and normal brain tissues in the TCGA, GETx and other three datasets from GlioVis. (**J, K**) UMAP plot depicting the distribution of ssGSEA scores among CD14^+^ monocytes between glioma samples and HCs (**I**). Boxplot compares the ssGSEA scores between two groups (**J**).

We next treated the 233 upregulated DEGs as a gene signature (SCMG, <u>S</u>ignature of <u>C</u>irculating <u>M</u>onocytes in <u>G</u>lioma) and calculated single-sample gene set enrichment analysis (ssGSEA) [22] scores for bulk RNA-seq data of peripheral blood monocytes from the GSE77043 cohort [21], which included four glioma patients and four HCs. As shown in **Figure 3E**, glioma patients exhibited significantly higher ssGSEA scores compared to HCs (Wilcoxon rank-sum test, *P* = 0.029), indicating consistent activation of the glioma-associated monocyte transcriptional program in an independent dataset. Similar findings were confirmed across multiple tissue-derived datasets (**Figures 3F-I**), suggesting that glioma-associated transcriptional signatures identified in circulating CD14^+^ monocytes may partially reflect tumor-intrinsic signals. Notably, when this gene set was projected back to the scRNA-seq data, glioma-derived circulating CD14^+^ monocytes also exhibit significantly elevated ssGSEA scores (Wilcoxon rank-sum test, *P* < 2.2e-16) (**Figures 3J, K**).

### Circulating CD14^+^ monocytes acquire glioma-specific transcriptional signatures

To investigate whether glioma-derived circulating CD14^+^ monocytes reflect glioma-intrinsic transcriptional features, we identified 557 glioma-specific genes that were significantly upregulated in gliomas compared to other cancer types using pan-cancer expression data from TCGA (**Supplementary Table S5**). Cross-referencing these 557 glioma-specific genes with the SCMG revealed 15 overlapping genes (**Figure 4A**, hypergeometric test, *P* = 2.98e-3). We next examined the expression of the 15 overlapping genes in circulating CD14^+^ monocytes from 79 glioma patients, 14 OBCs, and 33 OCs. Seven genes (SOX8 (*P* = 0.065), RGMA (*P* = 0.0006), GLDN (*P* = 0.046), RSPO2 (*P* = 0.011), LPL (*P* = 0.002), SLC1A3 (*P* = 0.073), and MYBPC1 (*P* = 0.003)) were upregulated in glioma-derived circulating CD14^+^ monocytes (**Figure 4B**). The expression profiles of these seven genes across TCGA pan-cancer datasets are presented in **Figure 4C**, exhibiting glioma-predominant overexpression patterns among different cancer types. The unsupervised clustering based on the SCMG revealed a clear separation between glioma samples from OBC or OC samples (**Figure 4D**). In addition, the GSEA analysis using the SCMG also confirmed these features were specifically associated with glioma (**Figures 4E, F**). These findings suggested that glioma-derived circulating CD14^+^ monocytes capture glioma-specific transcriptional features. By calculating ssGSEA scores for cancer hallmark pathways in each sample, we found that glioma-derived circulating CD14^+^ monocytes exhibited significant activation of “WNT/β-catenin signaling”, “MYC targets V2”, and “bile acid metabolism” pathways (**Supplementary Figure S4A**). These pathways’ activation may reflect tumor-driven transcriptional reprogramming in circulating CD14^+^ monocytes.

**Figure 4.**
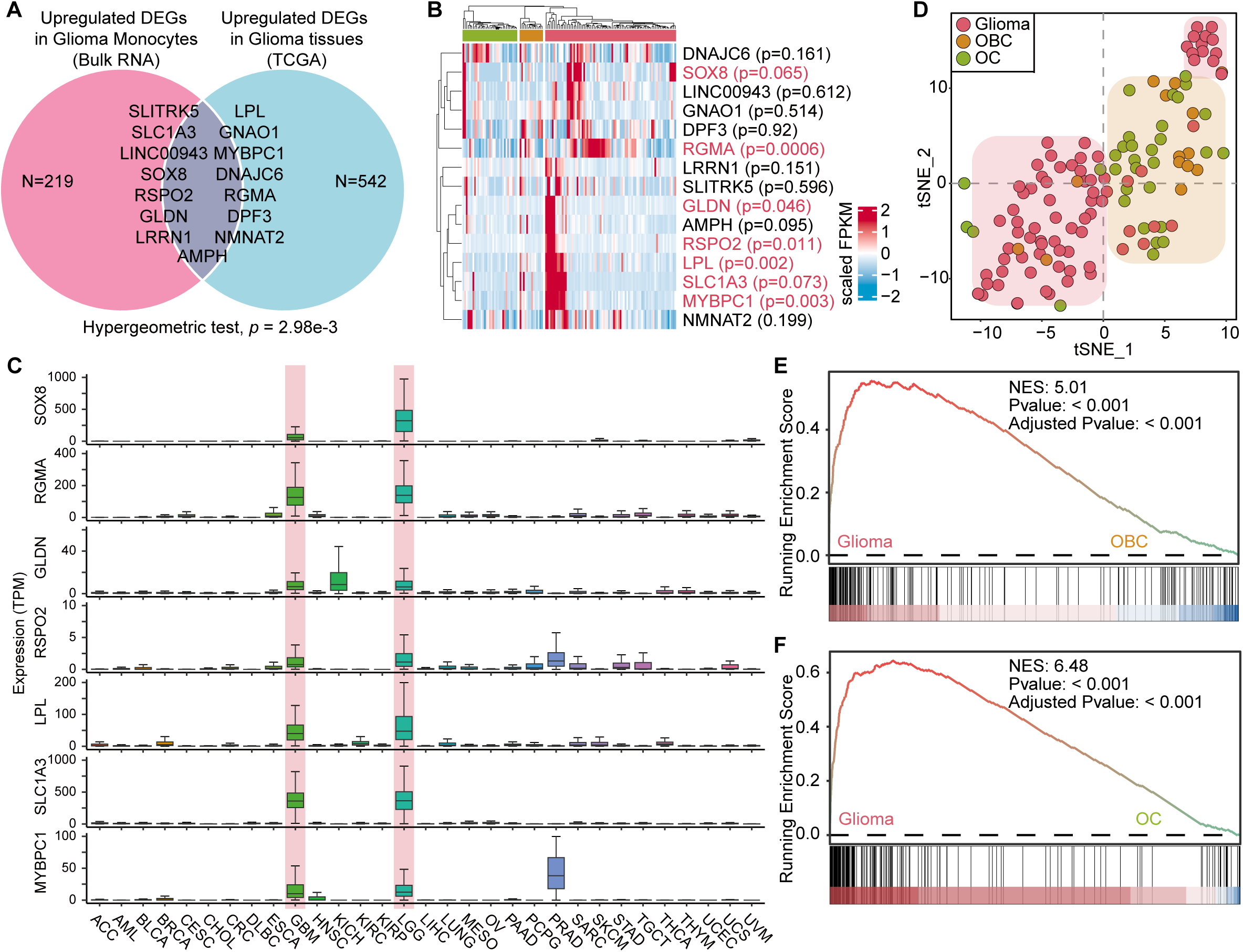
Bulk RNA-seq profiling of circulating CD14^+^ monocytes from glioma, OBC and OC samples. **(A)** Venn diagram showing the overlap between 233 upregulated DEGs from glioma-derived circulating CD14^+^ monocytes and 557 glioma-specific highly expressed genes identified from TCGA pan-cancer analysis. P value was calculated using the hypergeometric test. **(B)** The heatmap displays scaled expression profiles of 15 overlap genes across 79 glioma, 14 OBC and 33 OC samples, with rows corresponding to genes and columns to individual samples. P value was obtained by the Student’s t test. **(C)** Comparison of the expression levels of seven genes (SOX8, RGMA, GLDN, RSPO2, LPL, SLC1A3, and MYBPC1) across cancer types in the TCGA pan-cancer dataset. **(D)** t-SNE plot showing the result of unsupervised dimensionality reduction on 79 glioma, 14 OBC and 33 OC samples using the 233 upregulated DEGs from glioma-derived circulating CD14^+^ monocytes. OBC, other brain tumor; OC, other non-central nervous system cancers. (**E, F**) GSEA of the 233 upregulated DEGs was performed to compare glioma-derived circulating CD14^+^ monocytes with those from OBC (**E**) and OC (**F**) samples, respectively. Pathways with positive NES are enriched in glioma samples. FDR: false discovery rate; NES: normalized enrichment score.

### A diagnostic model for detection of glioma from expression profiling of circulating CD14^+^ monocytes

In light of the association between transcriptional features in circulating CD14^+^ monocytes and glioma, we next explored the potential of expression profiling of these monocytes as a sensitive approach for blood-based detection of gliomas. We applied a machine-learning approach in which a glioma diagnostic model was trained using a total of 690 DEGs found between gliomas and either other tumors or HCs (**Figures 5A, B and Supplementary Table S6**). An ensemble of GLMnet classifier was built by repeating the training procedure 50 times, using 75% of the samples each time. Each classifier was evaluated on its respective sub-training and sub-testing subsets, as well as on a holdout validation set, using receiver operating characteristic (ROC) analysis. The ensemble yielded robust performance across datasets: in sub-training subsets, the mean area under the ROC curve (AUC) was 0.999 (95% CI: 0.995–1.000; **Supplementary Figure S5A**); in sub-testing subsets, the mean AUC was 0.892 (95% CI: 0.686–1.000; **Supplementary Figure S5B**); and in the holdout validation set, the mean AUC was 0.936 (95% CI: 0.873–0.999; **Supplementary Figure S5C**). The median prediction score output by the ensemble classifier was defined as glioma risk score. We next applied the trained ensemble classifier to a holdout validation set comprising 31 samples, including glioma patients (n = 14), patients with all other tumors (n = 14), and HCs (n = 3) (**Supplementary Figure S5D** and **Figure 5C**). The ROC curves and corresponding AUC values illustrating the classifier’s performance in distinguishing gliomas from other tumors or HCs are shown in **Supplementary Figure S5E**.

**Figure 5.**
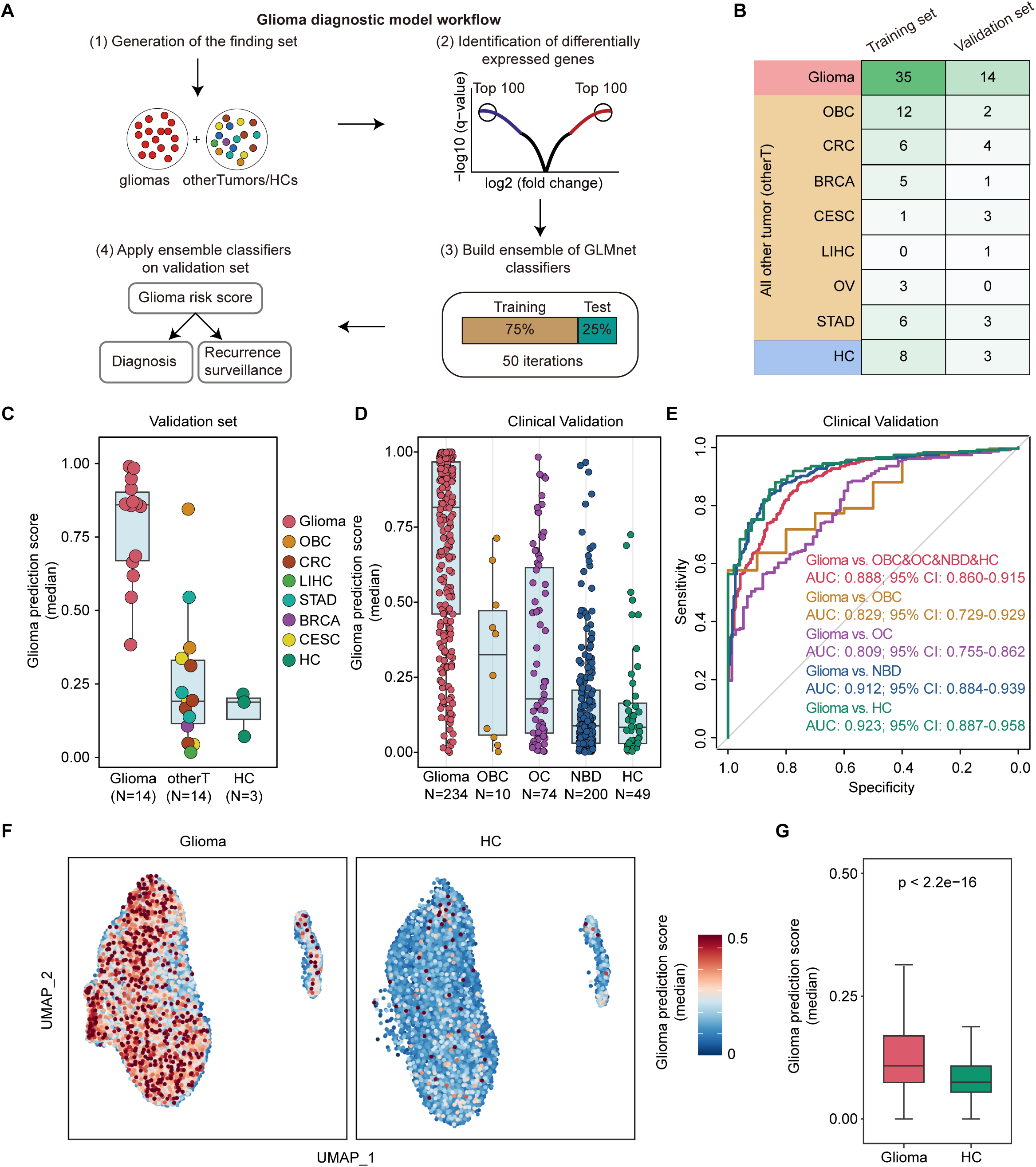
Generation of a classifier for blood-based detection of glioma. **(A)** Analysis workflow for the generation of the glioma diagnostic model. **(B)** Sample composition of the training set (n = 76) and holdout validation set (n = 31), showing the number of samples from each group. (**C, D**) Distribution of the median glioma prediction scores across different sample groups in the holdout validation set (C) and independent clinical validation set (D), respectively. **(E)** ROC curves and corresponding AUC values based on median glioma prediction scores for distinguishing glioma samples from OBC and HC samples in the independent clinical validation set. (**F, G**) UMAP plot depicting the distribution of median glioma prediction scores among CD14^+^ monocytes between glioma samples and HCs (**F**). Boxplot compares the median glioma prediction scores between two groups (**G**). P value was obtained by the Wilcoxon rank-sum test.

To further assess the generalizability of the ensemble classifier for glioma diagnosis, we applied the trained model to an independent clinical cohort, comprising 234 primary gliomas, 10 OBCs, 74 OCs, 200 NBDs, and 49 HCs (**Supplementary Table S2**). The distribution of glioma risk scores across all 567 independent validation samples is shown in **Figure 5D**. The classifier demonstrated robust glioma discrimination performance, achieving an AUC of 0.923 (95% CI: 0.887–0.958) for distinguishing gliomas from HCs, 0.912 (95% CI: 0.884–0.939) for gliomas versus NBDs, 0.809 (95% CI: 0.755–0.862) for gliomas versus OCs, 0.829 (95% CI: 0.729–0.929) for gliomas versus OBCs, and 0.888 (95% CI: 0.860–0.915) when comparing gliomas against OBCs, OCs, NBDs and HCs (**Figure 5E**). Given that the classifier was trained on expression profiles of circulating CD14^+^ monocytes, we further applied the trained model to CD14^+^ monocytes cluster extracted from scRNA-seq data. UMAP visualization illustrated distinct distributions of glioma risk scores between glioma patients and HCs (**Figure 5F**), with significantly higher scores observed in glioma-derived CD14^+^ monocytes (**Figure 5G**). These results underscore the potential of circulating CD14^+^ monocyte transcriptomic profiling as a noninvasive tool for glioma detection.

Furthermore, to explore the feasibility of constructing diagnostic models using molecular features beyond gene-level expression, we attempted to build a model based on exon-level expression profiles. Specifically, we applied the same machine learning pipeline as described above, but replaced the gene expression features with exon expression features. Using this approach, we identified and used 4,684 exon features (**Supplementary Table S7**) to train an ensemble classifier. This exon-based model also demonstrated promising classification performance in the independent clinical validation cohort (**Supplementary Figure S6A**). The classifier achieved robust discrimination, with a mean AUC of 0.977 (95% CI: 0.961–0.994) for distinguishing gliomas from HCs, 0.969 (95% CI: 0.953–0.986) for gliomas versus NBDs, 0.943 (95% CI: 0.919–0.968) for gliomas versus OCs, 0.892 (95% CI: 0.803–0.981) for gliomas versus OBCs, and 0.962 (95% CI: 0.945–0.980) when comparing gliomas against both OBCs, OCs, NBDs and HCs (**Supplementary Figure S6B**). These results suggested that exon-level expression features also hold diagnostic potential for glioma, providing an alternative molecular dimension for future model development and multi-feature integration.

### The glioma diagnostic model enables postoperative monitoring of glioma recurrence and prognosis evaluation

We subsequently investigated the potential use of the circulating CD14^+^ monocytes as an indicator of tumor recurrence. To this end, we applied the diagnostic model to 51 samples collected more than one year after surgery from glioma patients, including 28 patients without evidence of recurrence and 23 patients with radiologically confirmed recurrent glioma (**Figure 6A**). Notably, glioma risk scores were markedly elevated in recurrent cases compared to those without recurrence (**Figure 6B**). The diagnostic model demonstrated a high level of accuracy in distinguishing between these recurrence statuses, achieving an AUC of 0.975 (95% CI: 0.934–1.000) (**Figure 6C**). Furthermore, we compared paired intraoperative and one-week postoperative glioma prediction scores from 30 patients and observed a significant decline in scores following surgical resection (**Figures 6D, E**). Additionally, Kaplan–Meier analysis of postoperative glioma risk scores, categorized into low and high groups based on the median score, showed that patients with lower scores experienced significantly longer progression-free survival (PFS) compared to those with higher scores (**Figure 6F**; two-sided log-rank test, *P* = 0.034). In a multivariable Cox regression analysis, adjusting for age, extent of resection (EOR), and IDH mutation status, dichotomized postoperative glioma prediction scores remained associated with PFS, yielding a hazard ratio (HR) 11.68 (95% CI: 0.686–198.7; P = 0.0892) (**Figure 6G**). Overall, these findings indicate that expression profiling of circulating CD14^+^ monocytes holds potential clinical utility in glioma management, facilitating sensitive blood-based tumor detection and providing valuable insights into recurrence risk and prognosis.

**Figure 6.**
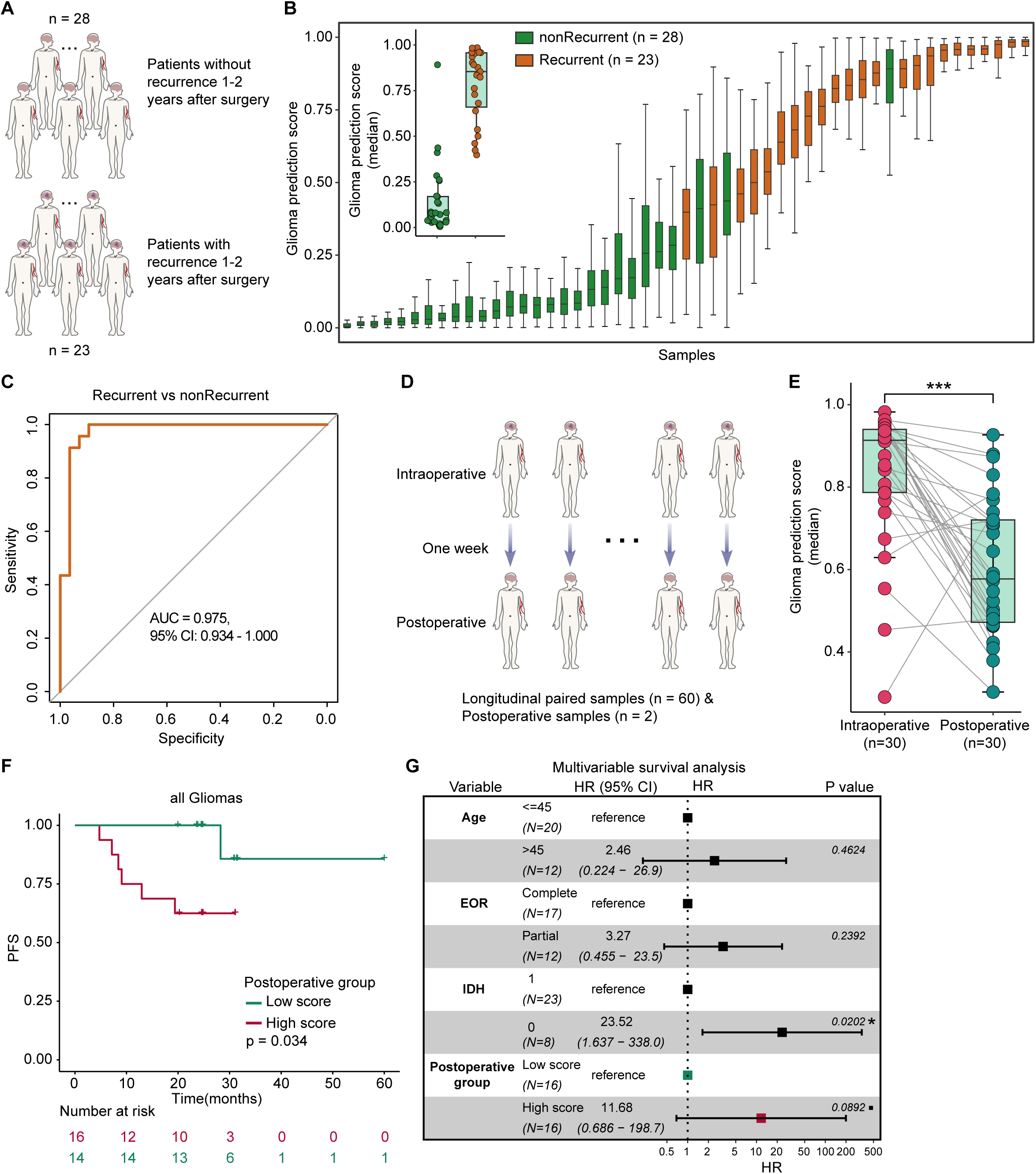
Glioma risk score implies recurrence and predicts survival in glioma patients. **(A)** Schematic overview of the state of glioma patients after surgery. **(B)** Box plots of classifier tumor prediction scores for 28 held-out nonrecurrent and 23 held-out recurrent glioma samples from applying the 50 classifiers. Boxes mark the 25th percentile (bottom), median (central bar) and 75th percentile (top); whiskers extend to minimum and maximum points. Inset plot shows the median glioma prediction scores for two groups of samples. **(C)** ROC curves and corresponding AUC values based on median glioma prediction scores for distinguishing 28 held-out nonrecurrent and 23 held-out recurrent glioma samples. **(D)** Schematic overview of the collecting of longitudinal paired samples. **(E)** Comparison of median glioma prediction scores in circulating CD14^+^ monocytes between matched intraoperative and one-week postoperative samples from 30 patients. P-value was calculated using the Wilcoxon signed-rank test. **(F)** Kaplan–Meier curves showing PFS of the 32 glioma patients stratified by high and low median glioma prediction scores (dichotomized using the median value). The number of patients at risk for each time point is indicated below the time point and color coded according to high or low groups. P value obtained by comparing the groups using a two-sided log-rank test. **(G)** Forest plot showing the results of multivariable Cox proportional hazards regression modeling of PFS for patients with median glioma prediction scores high or low status. Error bars indicate 95% CI for the HR. P values were calculated using a two-sided Wald test.

## Materials and methods

### Human Tumor and Peripheral Blood Specimens

This study was approved by the Ethics Committee of Nanjing Medical University (2017-391), Beijing Tiantan Hospital of Capital Medical University (KY2024-171-02), Nanjing First Hospital (KY20251021-01) and First Affiliated Hospital of Nanjing Medical University (2025-SR-975). The study was conducted in accordance with the Declaration of Helsinki and relevant ethical guidelines to ensure no potential risk to the participants. Peripheral blood specimens were sourced from four tertiary academic medical centers in China, including Beijing Tiantan Hospital (Beijing), Jiangsu Province Hospital (Nanjing), Tangdu Hospital of the Fourth Military Medical University (Xi’an), and Nanjing First Hospital (Nanjing). All patients were enrolled between July 2017 and January 2026. Peripheral blood specimens were sourced from these centers under institutional review board–approved protocols with written informed consent. Eligibility criteria required that all primary tumor patients were treatment free, meaning no prior surgery, radiotherapy, chemotherapy, or other anti tumor therapy, and that every tumor patient had a confirmed pathological diagnosis, while non neoplastic brain disorder (NBD) patients were included based on clinical diagnosis. Exclusion criteria consisted of glioma patients who also had other systemic malignancies or multiple malignancies, as well as any individuals whose blood volume was insufficient, whose sample quality was substandard, or whose samples could not be tested owing to errors during collection, extraction, storage, or experimental procedures.

Our discovery cohort comprised 209 samples (intraoperative blood collection unless otherwise specified) from 179 tumor patients, including: 79 primary gliomas, 30 matched and 2 unmatched post-resection samples (one week postoperatively), 23 recurrent gliomas, 28 recurrence-free gliomas (1-2 years postoperatively), 14 other brain tumors (OBC), 33 non-central nervous system cancers (OC); Eleven healthy donors served as healthy controls (HC) (**Supplementary Table S1**). For clinical validation, an independent cohort included 234 primary gliomas, 10 OBCs, 74 OCs, 200 non-neoplastic brain disorders (NBD) patients, and 49 HCs (**Supplementary Table S2**). Notably, longitudinal paired samples (intraoperative and one week postoperative) were secured from 30 treatment-naive glioma patients. These patients underwent 5 year survival follow up to assess progression free survival (PFS). All tumor patient outcomes were confirmed by clinical diagnosis combined with pathological gold standard criteria.

Fresh tumor tissues and blood samples were acquired from two IDH-wildtype glioblastoma patients (P0619 and P0708) during resection, which were subsequently used for single-cell RNA sequencing via 10x Genomics Chromium Single Cell 3’ Kit (v3.0 chemistry). All blood specimens were collected in K2E-EDTA Plus Blood Collection Tubes (BD, cat. no. REF 367525). Within 24 hours of phlebotomy, during which the samples were stored at 4°C, CD14^+^ cells were isolated.

### Sorting of CD14^+^ cells

Mononuclear cells were isolated by density gradient centrifugation using Ficoll-Paque™ (GE Life, cat. no. 17-1440-03), followed by washing with Dulbecco’s phosphate-buffered saline (DPBS; Biosharp, cat. no. BL310A). CD14^+^ monocytes were subsequently enriched by positive selection using anti-CD14 microbeads (Miltenyi Biotec, cat. no. 130-050-201; HUABIO, cat. no. HAK21014) according to the manufacturer’s protocol. Isolated CD14^+^ cells were immediately lysed and preserved in TRIzol™ reagent (Invitrogen, cat. no. 15596026CN) at -80°C until RNA extraction.

### RNA extraction and quantification

Total RNA was isolated using the VeZol-Pure Total RNA Isolation Kit (Vazyme, cat. no. RC202-01), following the manufacturer’s instructions. RNA concentration was determined using the Qubit™ RNA High Sensitivity (HS) Kit (Thermo Fisher Scientific, cat. no. Q32852) on a Qubit 4.0 Fluorometer, with purity assessed by A260/A280 and A260/A230 ratios measured via spectrophotometry (NanoDrop). RNA integrity was verified using Qseq1 Bio-Fragment Analyzer (BiOptic Inc.) with all samples.

### RNA Library Preparation and Sequencing

For 220 finding samples: VAHTS mRNA Capture Beads (Vazyme, cat. no. N401) was used for enriching mRNA from total RNA. RNA sequencing libraries were constructed using the VAHTS Universal V8 RNA-seq Library Prep Kit for Illumina (Vazyme, cat. no. NR605-01) following manufacturer-recommended protocols. The 220 finding libraries were sequenced on an Illumina HiSeq 2000 platform.

For 567 clinical validation samples: Ribo-off rRNA Depletion Kit (Vazyme, cat. no. N406-01) was used for removing ribosomal RNA (rRNA) from total RNA. RNA sequencing libraries were constructed using the VAHTS Universal V8 RNA-seq Library Prep Kit for Illumina (Vazyme, cat. no. NR605-01) following manufacturer-recommended protocols. The 567 libraries used for clinical validation were sequenced on Salus Pro sequencer.

### Single-Cell RNA Library Preparation and Sequencing

ScRNA-seq was performed using the 10x Genomics Chromium Single Cell 3’ Kit (v3.0 chemistry). CD14^+^ cell isolates were quantified using a Countess automated cell counter (Thermo Fisher Scientific) and adjusted to a concentration of 700-1,200 cells/µL prior to loading. Single-cell libraries were prepared following the manufacturer’s protocol and sequenced on an Illumina HiSeq X Ten system with 150-bp paired-end reads.

To expand the analytical scope, we collected raw scRNA-seq reads from public PBMC datasets available in the Gene Expression Omnibus (GEO) and Sequence Read Archive (SRA). These included GSE139324 [23, 24] (26 head and neck squamous cell carcinoma (HNSCC) patients and 6 healthy controls (HC)), GSE149689 [25, 26] and GSE264196 [27] (each comprising 4 HCs), GSE285888 [28] (33 non-small cell lung cancer (NSCLC) patients), and PRJNA883006 [29] (9 glioma specimens subjected to CD45^+^ immune cell enrichment).

### ScRNA-seq data generation and processing

The raw sequencing reads in FASTQ files were aligned to the human reference genome sequence (GRCh38.p13) using Cell Ranger (v3.1.0, 10X Genomic) software with default parameters to generate gene-barcode matrices. The Seurat (v4.4.0) R package [30] was applied for quality control, analysis, and exploration for all scRNA-seq data. For downstream analysis, only protein-coding genes were retained. Low-quality cells were excluded if they expressed fewer than 500 or more than 4,000 genes, or if over 15% of unique molecular identifiers (UMIs) linked to mitochondrial genes. Due to the large cell numbers in publicly available datasets, Harmony (v1.2.0) [31] was employed to integrate all public samples and further remove batch effects across individuals. Subsequent analyses employed Seurat’s pipeline, including neighborhood graph construction (“FindNeighbors”), and unsupervised clustering (“FindClusters”). For visualization, we reduced dimensionality using uniform manifold approximation and projection (UMAP) via “RunUMAP”, retaining the same principal components used for clustering. Cell identities were annotated based on established lineage-specific markers: T cells (CD2, CD3D, CD3E), NK cells (NKG7, KLRD1, GNLY), B cells (CD79A, MS4A1, CD40), Plasma cells (MZB1), CD14 Monocytes (CD14, S100A8, S100A9), CD16 Monocytes (FCGR3A, MS4A7), Dendritic cells (FCER1A, CST3) and Platelets (PPBP, PF4). We further identified subclusters for CD14^+^ monocytes of Gliomas and HCs by repeating the abovementioned steps, including harmony integration, dimensionality reduction, and clustering. Differentially expressed genes (DEGs) among CD14^+^ monocyte subpopulations were identified using “FindAllMarkers”. Genes with the criteria of an adjusted p value < 0.05 and average log2 fold change (FC) > 0.25 were defined as DEGs.

### Cell differentiation analysis

To explore the varying degrees of differentiation among distinct CD14^+^ monocytes of Gliomas and HCs, we employed the R package CytoTRACE2 (v1.0.0) [20] to obtain the CytoTRACE2 score for each cell, enabling to infer their respective differentiation states. The CytoTRACE2 scores ranging from 0 to 1 were assigned, with higher scores indicative of heightened stemness (lesser differentiation), and conversely, lower scores reflecting lower stemness.

### RNA-seq data generation and processing

The raw sequencing reads in FASTQ files were cleaned using fastp (v0.23.4), including adapter trimming and removal of low-quality reads. Filtered reads were mapped to the human reference genome (GRCh38.p13) using STAR aligner (v2.7.7a) with default splicing-aware parameters. Gene-level counts were generated via htseq-count (v2.0.3) in union mode against GCA_000001405.28 annotations. Raw counts were normalized to FPKM values using the formula:

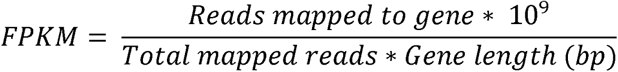

The final gene expression matrix was rigorously subset to exclusively retain protein-coding genes prior to downstream analyses.

### Differential expression and Pathway enrichment analysis

The significantly DEGs between glioma samples and healthy controls were identified using the edgeR (v3.42.4) package [32]. Genes with the criteria of an adjusted p value < 0.1 and FC > 2 were defined as DEGs. For functional inference of DEGs, the Enrichr(https://maayanlab.cloud/Enrichr/) were used for gene functional enrichment analysis, and the KEGG pathways were selected as background gene sets. Gene set enrichment analysis (GSEA) was performed using the clusterProfiler (v4.8.3) package [33] to assess the enrichment of cancer hallmark pathways among DEGs between glioma samples and healthy controls. Additionally, pathways or gene sets activity scores were computed using the single sample gene set enrichment analysis (ssGSEA) from GSVA (v1.48.3) R package [34].

### Construction of glioma diagnostic model

To establish a dedicated glioma diagnostic framework, we defined non-glioma controls as individuals with other intracranial tumors, extra-cranial malignancies, or healthy status. A discovery cohort of 107 treatment-naive primary glioma patients was curated by excluding 60 paired intra-/post-operative samples and 51 previously treated cases. This cohort was stratified 7:3 into training (n = 76) and holdout validation (n = 31) sets. Through 50 bootstrap iterations, the training set was further partitioned 3:1 into sub-training (n = 57) and sub-testing (n = 19) subsets, in which edgeR was used to identify the top 100 up- and down-regulated genes. These features were then used to train GLMnet classifiers via the caret framework (v6.0.94), employing repeated 5-fold cross-validation (3 repeats) and elastic net hyperparameter tuning (α = 0-1, λ = 0-0.1). Each classifier was evaluated on its corresponding test subset using ROC analysis. The final ensemble-derived glioma risk score, applied to the holdout validation set, was calculated as the median predicted probability across all 50 models. Preclinical validation was subsequently conducted in an independent cohort (n = 567), evaluating the clinical utility.

### Glioma risk score and survival analysis

Patients with primary glioma were stratified into high- and low-risk groups based on the median glioma risk score measured one week postoperatively (n = 32), to evaluate prognostic differences. Survival analysis was performed using the R package survival (v3.5.5), and Kaplan–Meier curves were generated with the ggsurvplot function from the survminer package (v0.4.9). Multivariable Cox proportional hazards regression models were fitted using the glioma risk score as either a dichotomized or continuous variable, adjusting for age, extent of resection (EOR), and IDH mutation status. Results were visualized using the ggforest function from the survminer package.

### Statistics and reproducibility

No statistical methods were used to predetermine sample sizes, and no data were excluded from the analyses. Prior to model construction, investigators were blinded during the stratification of the 107 discovery cohort samples, which were divided into training and validation sets in a 7:3 ratio, and no subsequent reallocation was performed. No imputation or exclusion was performed for missing clinical variables (age, IDH status, EOR). RNA-seq-derived predictor variables had no missing data. ScRNA-seq data were analyzed using R (v4.1.3), while bulk RNA-seq data were analyzed with R (v4.3.1). This study is reported as per the Transparent Reporting of a Multivariable Prediction Model for Individual Prognosis or Diagnosis (TRIPOD) guideline (Supplementary File S1 Checklist).

## Discussion

Glioma, particularly GBM, remains one of the most devastating central nervous system malignancies, with a median survival of less than 15 months despite aggressive treatment [1, 2]. Accurate diagnosis and early detection of recurrence are critical but remain major clinical challenges [35, 36]. Traditional imaging tools such as MRI are often insufficient to distinguish true progression from pseudo progression and lack the sensitivity to detect early-stage tumors or minimal residual disease [37, 38]. Tissue biopsy, while informative, is invasive and subject to spatial sampling errors due to the heterogeneous nature of gliomas [39–41].

In this study, we investigated circulating CD14^+^ monocytes as a potential biomarker source for glioma diagnosis and recurrence monitoring. Several previous studies have suggested that TAMs in glioma originate primarily from peripheral blood monocytes [17–19]. These monocytes are recruited into the tumor microenvironment, where they acquire immunosuppressive and tumor-promoting functions. Our findings support and extend this concept by showing that circulating CD14^+^ monocytes in glioma patients exhibit abnormal proportions, impaired differentiation, and transcriptional profiles indicative of tumor influence.

We observed a significant elevation in CD14^+^ monocyte abundance in the peripheral blood of glioma patients compared to healthy controls. Importantly, these monocytes exhibited global transcriptional dysregulation, including upregulation of hallmark cancer pathways such as PI3K-Akt signaling, epithelial–mesenchymal transition, and inflammatory response. CytoTRACE2 analysis confirmed that these cells had reduced differentiation status, reflecting a shift toward a more progenitor-like phenotype, possibly driven by tumor-derived signals. These findings align with prior research identifying glioma-induced systemic immune reprogramming. By integrating data from scRNA-seq and bulk RNA-seq platforms, we identified a set of 233 upregulated genes in glioma-derived CD14^+^ monocytes, some of which overlapped with tumor-intrinsic glioma markers derived from pan-cancer TCGA analysis. Notably, seven genes (SOX8, RGMA, GLDN, RSPO2, LPL, SLC1A3, and MYBPC1) were consistently overexpressed in glioma monocytes and in glioma tissue, confirming the potential of these cells as surrogates of tumor biology. To translate these insights into a clinically actionable tool, we developed an ensemble diagnostic model using 50 GLMnet classifiers trained on the expression profiles of 690 DEGs. The model demonstrated strong discriminative power across multiple datasets, achieving AUCs about 0.9 in both discovery and independent validation cohorts. Notably, the model retained high specificity in distinguishing glioma from other central and non-central nervous system cancers, underscoring the specificity of the monocyte-derived signature.

Beyond diagnosis, we evaluated the utility of our model for recurrence monitoring. In a longitudinal cohort, glioma prediction scores significantly declined post-surgery and were elevated again in cases with radiologically confirmed recurrence. Moreover, high postoperative prediction scores were associated with shorter progression-free survival, even after adjusting for confounding clinical variables such as age, extent of resection, and IDH mutation status, suggesting that circulating monocytes can reflect residual disease and may serve as a liquid biomarker for postoperative surveillance and risk stratification.

Compared with existing diagnostic modalities, our monocyte-based approach offers a noninvasive way which is compatible with routine blood draws, facilitating serial monitoring. It captures dynamic and systemic immune responses that may precede radiographic changes. Moreover, it provides a molecular readout that complements imaging-based and histological assessments, especially in ambiguous cases such as pseudo progression.

While our study reveals systemic transcriptional reprogramming of circulating monocytes in glioma, the mechanisms by which tumors influence these cells and the pathways giving rise to their glioma-specific transcriptional features remain poorly understood. We hypothesize that tumor-derived extracellular vesicles or soluble factors may traverse the BBB and reshape monocyte transcriptional programming. Future studies employing bone marrow chimera models and spatial multi-omics could delineate whether these changes require continuous tumor crosstalk or become imprinted as a stable ‘tumor-educated’ state. Such insights may unlock novel immunomodulatory strategies for glioma management. In addition, the discovery and validation cohorts, although sizeable, were from a limited number of centers, and additional multicenter studies are warranted to confirm the generalizability of the findings. The classifier’s robustness across diverse platforms and preprocessing pipelines also requires further evaluation. Additionally, while we focused on CD14^+^ monocytes, other circulating immune subsets, such as T cells or dendritic cells, may contribute additional diagnostic information.

In summary, we provide compelling evidence that glioma induces systemic transcriptional reprogramming of circulating CD14^+^ monocytes, which can be leveraged for noninvasive diagnosis and postoperative monitoring. This immune-centered strategy offers a scalable and clinically feasible approach that may complement current imaging and tissue-based diagnostics, and opens new avenues for immune monitoring in neuro-oncology.

## Data availability

All public datasets used in this study are available in the Gene Expression Omnibus (https://www.ncbi.nlm.nih.gov/geo/) and the UCSC Xena (https://xenabrowser.net/datapages/). The processed gene expression profiles and any additional information required to reanalyze the data reported in this paper are available from the lead contact upon reasonable request. The ensemble machine learning diagnostic model developed in this study could be available from the GitHub repository https://github.com/PoShine/Glioma-Monocytes/tree/main.

## Supplementary Information

Supplementary Data are available at supplementary files.

## Acknowledgements

This work was supported by the Noncommunicable Chronic Diseases-National Science and Technology Major Project (2024ZD0525300), the National Natural Science Foundation of China (Grant Nos. 82372897, 82272867, 82503866, 91959113), China Postdoctoral Science Foundation (Grant No. 2025M772304), Beijing Natural Science Foundation (Grant No. 7254340), Beijing Municipal Science and Technology Commission (Z241100009024044), and Fundings of Beijing Neurosurgical Institute (11000025T000003319495). The authors acknowledge the technical support from all CGGA member for their help in clinical sample collection, and support from Jiangsu Brainovel Medical Technology Co., Ltd., specifically for high-throughput sequencing and data analysis. A select number of illustrations were drawn using BioRender. Special thanks to ChatGPT for enhancing the readability and language flow of this manuscript. ChatGPT did not replace any researcher tasks, including generating scientific insights, analyzing and interpreting data, or drawing scientific conclusions.

## Author contributions

Q.H.W. and T.J. designed and supervised the project. Q.H.W. secured the funding. W.W. performed the experiments and generated the data. X.P. analyzed the data and wrote the manuscript. W.W., R.C.C., and L.X.W. edited the manuscript. R.C.C., B.P., X.Y.C. and B.Q.Y. assisted with clinical database management and biospecimen coordination. D.C., Y.W., N.W., X.J.L., H.G.L., Q.W.D., F.W., F.L., L.W., W.Z., J.X.Z., and T.J. provided clinical samples. Q.H.W., T.J., J.X.Z., W.Z., L.W., and R.C.C. directed the research and contributed to the final version of the manuscript. All authors reviewed the manuscript.

## Competing interests

All authors declare that they have no competing interests.

**Supplementary Figure S1. Basic clustering information of 16 clusters in 82 samples.** (A) The UMAP plots showing 16 cell clusters identified by integrated analysis, along with the distribution of cell origins across different tissue types. Each dot corresponds to a single cell.

(B) Dot plots showing average expression of known markers in 16 cell clusters. The dot size represents percent of cells expressing the genes in each cluster. The expression intensity of markers is shown.

**Supplementary Figure S2. Analysis of peripheral blood CD14^+^ sorted cells collected from two glioma patients.** (A) The UMAP plots showing 10 cell clusters identified by integrated analysis, along with the distribution of cell origins across samples. Each dot corresponds to a single cell.

(B) Dot plots showing average expression of known markers in 10 cell clusters. The dot size represents percent of cells expressing the genes in each cluster. The expression intensity of markers is shown.

(C) UMAP plot showing major 5 cell types of 2 samples identified by integrated analysis. Each dot corresponds to a single cell, colored by cell types. NK cells, natural killer cells.

**Supplementary Figure S3. Differential expression analysis of circulating CD14^+^ monocytes from glioma samples and HCs.** (**C)** Differential gene expression analysis of circulating CD14^+^ monocytes between glioma samples and HCs. Colored DEGs meet the criteria of an adjusted p-value < 0.1 and fold change > 2.

(**D**) The heatmap displays scaled expression profiles of 258 DEGs across 79 glioma samples and 11 HCs, with rows corresponding to genes and columns to individual samples. Associated clinicopathological features are annotated above the heatmap columns.

**Supplementary Figure S4. Characterization of activated cancer hallmark pathways in circulating CD14^+^ monocytes across different types of cancer samples.** (**A**) ssGSEA was performed to evaluate the activation levels of 50 cancer hallmarks across three groups of samples: 79 glioma, 14 OBC, and 33 OC samples. The 50 cancer hallmarks were categorized into eight major classes: Signaling, Proliferation, Immune, Development, Metabolic, Cellular Component, DNA Damage, and Pathway. P value was obtained by the Wilcoxon rank-sum test; ***, p < 0.001; **, p < 0.01; *, p < 0.05; _·_, p < 0.1; ns, p ≥ 0.1.

**Supplementary Figure S5. Performance evaluation of 50 individual models within the ensemble classifier.** (**A-C**) ROC curves illustrating the performance of each model iteration in distinguishing glioma from non-glioma samples across the sub-training (A), sub-testing (B), and holdout validation (C) sets. The average AUC of the 50 models was calculated for each set.

(**D**) Box plots of classifier tumor prediction scores for the holdout validation set from applying the 50 classifiers. Boxes mark the 25th percentile (bottom), median (central bar) and 75th percentile (top); whiskers extend to minimum and maximum points.

(**E**) ROC curves and corresponding AUC values based on median glioma prediction scores for distinguishing glioma samples from other tumor and HC samples in the validation set.

**Supplementary Figure S6. Validation of the model based on exon-level expression profiles.** (A) Distribution of the median glioma prediction scores across different sample groups in the independent clinical validation set, respectively.

(E) ROC curves and corresponding AUC values based on median glioma prediction scores for distinguishing glioma samples from OBC, OC, NBD, and HC samples in the independent clinical validation set.

**Supplementary Table S1.** Summary of clinical characteristics and next generation sequencing (NGS) metrics for the 220 samples.

**Supplementary Table S2.** Summary of clinical characteristics and next generation sequencing (NGS) metrics for the 567 samples from the independent clinical validation set.

**Supplementary Table S3.** DEGs identified by comparing Mono_C0 CD14^+^ monocyte subcluster against the other three subclusters, respectively.

**Supplementary Table S4.** DEGs identified by comparing the gene expression profiles of circulating CD14^+^ monocytes from 79 glioma patients and 11 HCs.

**Supplementary Table S5.** A total of 557 glioma-upregulated DEGs identified by comparing glioma with 29 other cancer types in the TCGA pan-cancer gene expression dataset.

**Supplementary Table S6.** The 690 feature genes employed in the ensemble classifier, along with the number of individual classifiers (out of 50) in which each gene was utilized.

**Supplementary Table S7.** Detailed information of the 4,684 exon features.

